# Prehospitalization Proton Pump Inhibitor (PPI) use and Clinical Outcomes in COVID-19

**DOI:** 10.1101/2020.07.12.20151084

**Authors:** Preethi Ramachandran, Abhilash Perisetti, Mahesh Gajendran, Farla Jean-Louis, Pardeep Bansal, Alok Kumar Dwivedi, Hemant Goyal

**Author notes:** **Corresponding author:** Hemant Goyal, MD FACP PGDCA (MBA), The Wright Center for Graduate Medical Education, 501 S. Washington Avenue, Scranton, PA 18505. Preethi Ramachandran and Abhilash Perisetti share equal authorship. **Author contribution** Conception and design: Hemant Goyal. Formal Analysis: Mahesh Gajendran, Preethi Ramachandran. Literature search: Abhilash Perisetti, Mahesh Gajendran, Hemant Goyal. First draft: Abhilash Perisetti, Mahesh Gajendran, Preethi Ramachandran. Critical revision and editing: All authors. Final approval: All authors.

## Abstract

**Background and Aim:** Gastric acid has shown to neutralize many viruses. The working receptor of SARS-CoV-2 is angiotensin-converting enzyme-2 (ACE-2), which has shown to be omnipresent in the gastrointestinal tract. There is a theoretical concern that SARS-CoV-2 can escape the neutralization by gastric acid because of hypochlorhydria caused by the use of proton pump inhibitors (PPI) and can predispose the patients for severe COVID-19.

**Methods:** We studied the association between prehospitalization PPI use and clinical outcomes among hospitalized COVID-19 patients.

**Results:** In our study, 15.6% of hospitalized COVID-19 patients were on PPIs at home. Mortality among PPI-users was 2.3 times higher than non-users, along with 2.5 times higher risk of mechanical ventilation. This relationship existed even after adjusting for confounding variables.

**Conclusion:** These results warrant further investigation in prospective studies to evaluate if PPI-induced hypochlorhydria is associated with worse outcomes, including mortality because of the omnipresence of ACE-2 in the gastrointestinal tract.

## Introduction

The Coronavirus Disease-2019 (COVID-19) is caused by severe acute respiratory syndrome coronavirus-2 (SARS-CoV-2) [1, 2]. Emerging evidence suggests that COVID-19 can also present with gastrointestinal (GI) symptoms, including nausea, vomiting, dysgeusia, diarrhea, and abdominal pain [2, 3, 4, 5, 6]. The Angiotensin-converting enzyme 2 (ACE2) has been identified as the receptor for the SARS-CoV-2 to enter the epithelial cells, and it is highly expressed throughout the GI system, including the gastric epithelium [6]. It is thought that gastric acid could neutralize the virus on ingestion, and there is a concern that decreased gastric acid secretion by proton pump inhibitors (PPI) could potentially increase the risk of GI symptoms and might increase severity in COVID-19 [7, 8]. Moreover, previous studies have shown that there is an increased risk of infections and all-cause mortality in patients with prolonged use of PPIs [9]. However, no study has determined an association between the history of PPI use and clinical outcomes, particularly mortality in COVID-19. We sought to evaluate the association between PPI and clinical outcomes among hospitalized COVID-19 patients.

## Methods

It is a retrospective cohort study conducted on patients admitted with a principal diagnosis of COVID-19 in a tertiary care academic medical center in Brooklyn, New York, from March 1^st^ to April 25^th^, 2020. Patients were excluded if they were younger than 18 years, who were not hospitalized and managed on an ambulatory basis, pregnant, unavailability of the results of SARS-CoV-2 nasopharyngeal testing, and missing data on mortality or disposition. Data related to patients’ demographics, clinical symptoms, comorbidities, history of medications, vitals at presentation, admission laboratory tests, inpatient medications, and outcomes were collected.

In our study, the primary exposure of interest was the history of PPI use as a home medication. The home PPI use was determined based on the documentation of any type of PPI use (including omeprazole, esomeprazole, rabeprazole, pantoprazole, etc.) as an active home medication at the time of admission. The study cohort was divided into two groups based on PPI use at home: COVID-19 patients on PPI (cases) and COVID-19 patients not on PPI (controls). The primary outcomes are all-cause mortality and mechanical ventilation. The secondary outcomes are septic shock and hospital length of stay.

Statistical analyses were performed using IBM SPSS software version 26 (SPSS Inc, Armonk, NY) and STATA 15.1 (StataCorp LLC, College Station, TX). Two models of multivariable logistic regression analysis were conducted for evaluating the effect of PPI use on the primary dichotomous outcome variables, death, and mechanical ventilation separately. In Model 1, we adjusted for any significant variables associated with PPI (P<0.05) based on the univariate analysis. In Model 2, we performed the multivariable analysis based on clinically relevant confounders associated with outcome and PPI use (BMI, Age, HTN, Hyperlipidemia, CAD, DM, Cancer, COPD, aspirin, statin, smoker, gender, H2 blocker use). All the non-significant predictors were dropped from the final model and retained the maximum five variables to avoid any over-fitting issue in the multivariable model. The effect size of PPI use obtained from logistic regression analyses was summarized using the odds ratio (OR) along with a 95% confidence interval (CI). P-values less than or equal to 5% were considered statistically significant results. We also performed subgroup analyses, according to essential cofactors, to examine the heterogeneity of PPI effects on mortality outcomes (Forest Plot).

The study was approved by the Institutional Review Board (Protocol 20-12) of Brookdale University.

## Results

A total of 295 hospitalized COVID-19 patients formed our final study population. Of these, 15.6% (46/295) patients had documentation of any PPI use before admission (Figure 1). Baseline demographics, laboratory, and outcomes data are shown in table 1. We observed increased mortality in patients with a history of PPI compared to patients without PPI (34.8% vs. 16.2%, P=0.007). The odds of mortality remained significantly higher in the PPI group compared to control (OR 2.3, 95% CI 1.1 – 4.8, P=0.03) even after adjusting for potential factors in multivariate analysis (Table 2).

**Table 1:** Baseline characteristics, laboratory findings and clinical outcomes of the study population ^€^

| Characteristic | PPI Group<br>N=46<br>N (%) | No-PPI group<br>N=249<br>N (%) | P-value |
| --- | --- | --- | --- |
| **Baseline Demographics** |  |  |  |
| Age, median (IQR) | 67 (57.3, 76.5) | 65 (54, 74) | 0.38 |
| Age > 60 years | 32 (69.6) | 164 (65.9) | 0.74 |
| Female gender | 21 (45.7) | 112 (45) | 0.91 |
| BMI, mean (SD) | 27.3 (25.3, 32.4) | 29.3 (26.3, 34.9) | 0.008 |
| Race |  |  | 0.94 |
| • White | 2 (4.3) | 7 (2.8) |  |
| • African American | 33 (71.7) | 175 (70.3) |  |
| • Hispanic | 5 (10.9) | 34 (13.7) |  |
| • Asian | 2 (4.3) | 8 (3.2) |  |
| • Unknown | 4 (8.7) | 25 (10) |  |
| Charlson's Comorbidity Index |  |  | 0.15 |
| • Zero | 3 (6.5) | 44 (17.7) |  |
| • One | 9 (19.6) | 34 (13.7) |  |
| • Two | 10 (21.7) | 67 (26.9) |  |
| • Three or more | 24 (52.2) | 104 (41.8) |  |
| Comorbidities |  |  |  |
| • Hypertension | 39 (84.8) | 170 (68.3) | 0.02 |
| • Dyslipidemia | 24 (52.2) | 73 (29.3) | 0.004 |
| • CAD | 14 (30.4) | 31 (12.4) | 0.004 |
| • DM | 27 (58.7) | 105 (42.2) | 0.052 |
| • Cancer | 6 (13) | 14 (5.6) | 0.1 |
| • COPD | 10 (21.7) | 17 (6.8) | 0.004 |
| • Asthma | 7 (15.2) | 37 (14.9) | 1 |
| Immunocompromised Status | 5 (10.9) | 19 (7.6) | 0.56 |
| Smoker | 9 (19.6) | 23 (9.5) | 0.07 |
| Medications |  |  |  |
| • ACEI/ARB | 17 (37) | 76 (30.5) | 0.61 |
| • NSAID | 10 (21.7) | 35 (14.6) | 0.27 |
| • Aspirin | 22 (47.8) | 66 (27.5) | 0.009 |
| • Statin | 33 (71.7) | 81 (33.8) | <0.001 |
| Symptoms |  |  |  |
| Cough | 24 (52.2) | 154 (61.8) | 0.25 |
| • Fever | 24 (52.2) | 153 (61.4) | 0.47 |
| • Dyspnea | 28 (60.9) | 174 (69.9) | 0.23 |
| • Fatigue | 15 (32.6) | 96 (38.6) | 0.51 |
| • Myalgia | 10 (21.7) | 68 (27.3) | 0.47 |
| • GI symptom | 7 (15.2) | 46 (18.5) | 0.68 |
| **Laboratory Findings** |  |  |  |
| Anemia | 24 (52.2) | 110 (44.4) | 0.34 |
| Leukocytosis | 10 (21.7) | 61 (24.6) | 0.85 |
| Lymphopenia | 21 (45.7) | 148 (59.7) | 0.1 |
| Thrombocytopenia | 12 (26.1) | 49 (19.8) | 0.33 |
| Elevated Creatinine | 25 (54.3) | 85 (34.6) | 0.01 |
| Hypoalbuminemia | 14 (30.4) | 66 (26.6) | 0.59 |
| Elevated Lactate* | 21 (53.8) | 76 (38) | 0.08 |
| Elevated LDH* | 37 (100) | 215 (99.5) |  |
| Elevated CRP* | 15 (45.5) | 111 (54.1) | 0.45 |
| Elevated AST | 11 (36.7) | 47 (40.9) | 0.84 |
| Elevated ALT | 5 (16.7) | 16 (13.9) | 0.77 |
| Elevated Bilirubin | 3 (10) | 9 (7.8) | 0.71 |
| Elevated Alkaline Phosphatase | 7 (23.3) | 13 (11.3) | 0.13 |
| **Clinical Outcomes** |  |  |  |
| Shock | 16 (34.8) | 77 (30.9) | 0.61 |
| Mechanical Ventilation | 12 (26.1) | 37 (14.9) | 0.16 |
| Mortality | 16 (34.8) | 40 (16.2) | 0.007 |
| Length of stay, median (IQR) | 7 (4,10) | 6 (4,10) | 0.48 |

€ Non-parametric test (Mann-Whitney test) used for non-normal distributed continuous variable; SD- standard deviation; IQR- interquartile range \* Lactate 14% missing values; LDH 19% missing values; CRP 19% missing values

**Table 2:** Adjusted effects of PPI use on mortality and mechanical ventilation outcomes

| In-hospital Mortality |  |  |  |  |
| --- | --- | --- | --- | --- |
|  | Odds Ratio | 95% Confidence Interval |  | P-value |
| Multivariable Analysis Model 1 |  |  |  |  |
| Home PPI use | 2.24 | 1.07 | 4.71 | 0.033 |
| CCI | 1.01 | 0.82 | 1.24 | 0.921 |
| BMI | 1.01 | 0.97 | 1.05 | 0.547 |
| Home statin use | 1.59 | 0.84 | 3.02 | 0.157 |
| Multivariable Analysis Model 2 |  |  |  |  |
| Home PPI use | 2.33 | 1.13 | 4.83 | 0.022 |
| Diabetes Mellitus | 2.15 | 1.15 | 4.01 | 0.016 |
| Cancer | 4.1 | 1.52 | 11.09 | 0.005 |
| Female gender | 0.56 | 0.3 | 1.06 | 0.075 |
| Mechanical Ventilation |  |  |  |  |
| Multivariable Analysis Model 1 |  |  |  |  |
| Home PPI use | 2.21 | 1.00 | 4.90 | 0.051 |
| BMI | 1.04 | 1.00 | 1.08 | 0.063 |
| CAD | 2.62 | 1.11 | 6.21 | 0.028 |
| CCI | 0.79 | 0.63 | 1.00 | 0.055 |
| Multivariable Analysis Model 2 |  |  |  |  |
| Home PPI use | 2.49 | 1.14 | 5.43 | 0.022 |
| BMI | 1.06 | 1.02 | 1.1 | 0.004 |
| Female gender | 0.51 | 0.26 | 1.01 | 0.053 |

PPI: Proton Pump Inhibitor; BMI: Body Mass Index; CAD: Coronary Artery Disease; CCI: Charlson's comorbidity Index; IPW: inverse probability weighting; Model 1 adjusted only important variables associated with the outcome; Model 2 adjusted for all significant confounders associated with outcome.

**Figure 1:**
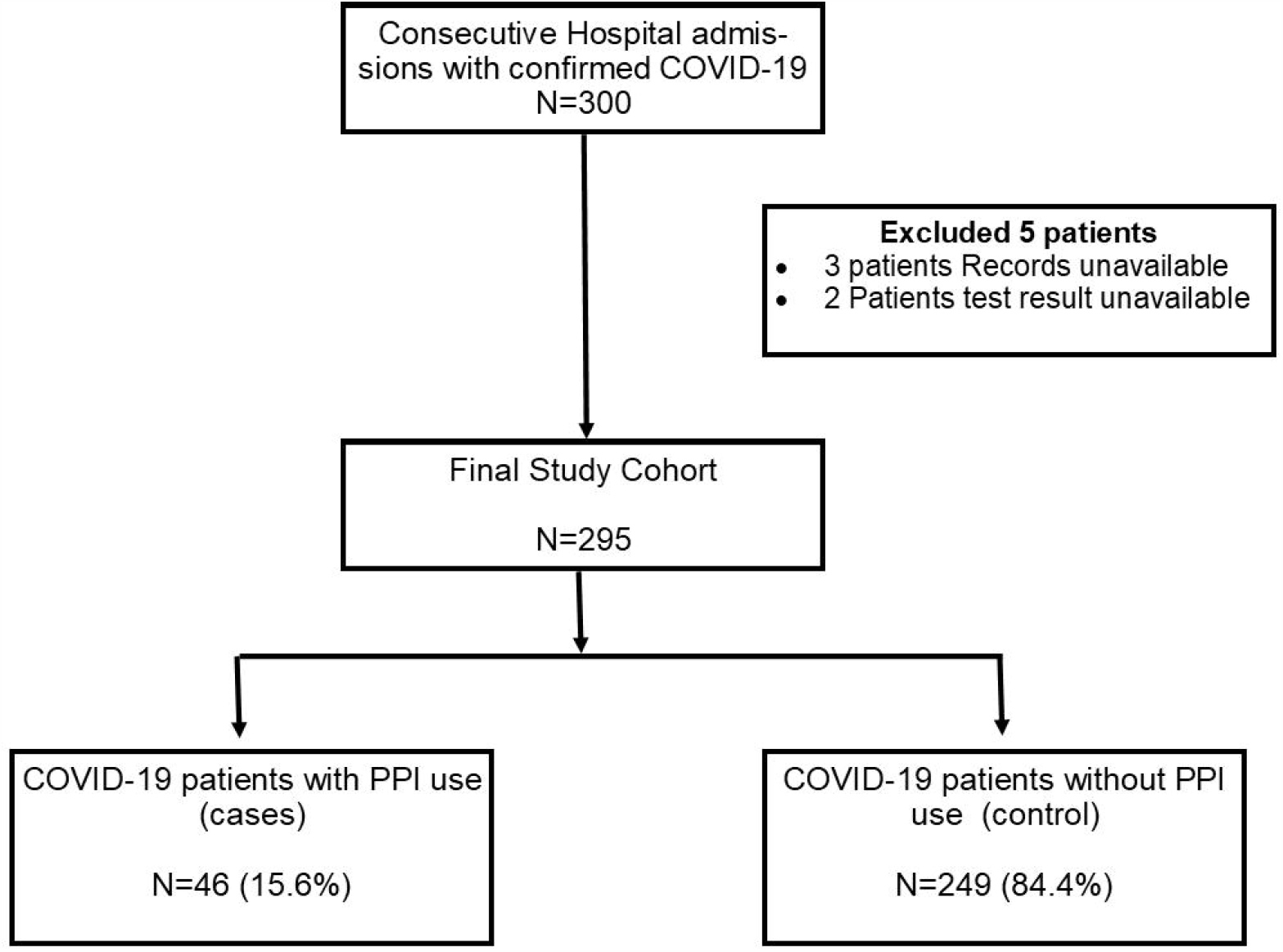
Study flow diagram.

Furthermore, the odds of mechanical ventilation were also observed to be higher in cases when compared to controls after adjusting for potential confounders as well as predictors (OR 2.5, 95% CI 1.1–5.4, P =0.02). The PPI use was associated with an increased odds of mortality in most subgroups of patients, as shown in the Forest Plot (Figure 2). The profound effect of PPI on mortality was observed in younger patients without cardiac risk factors (diabetes, hypertension, dyslipidemia, CAD) or Charlson’s comorbidity index of zero, suggesting that a history of PPI exacerbate prognosis even in patients without having underlying cardiac conditions.

**Figure 2:**
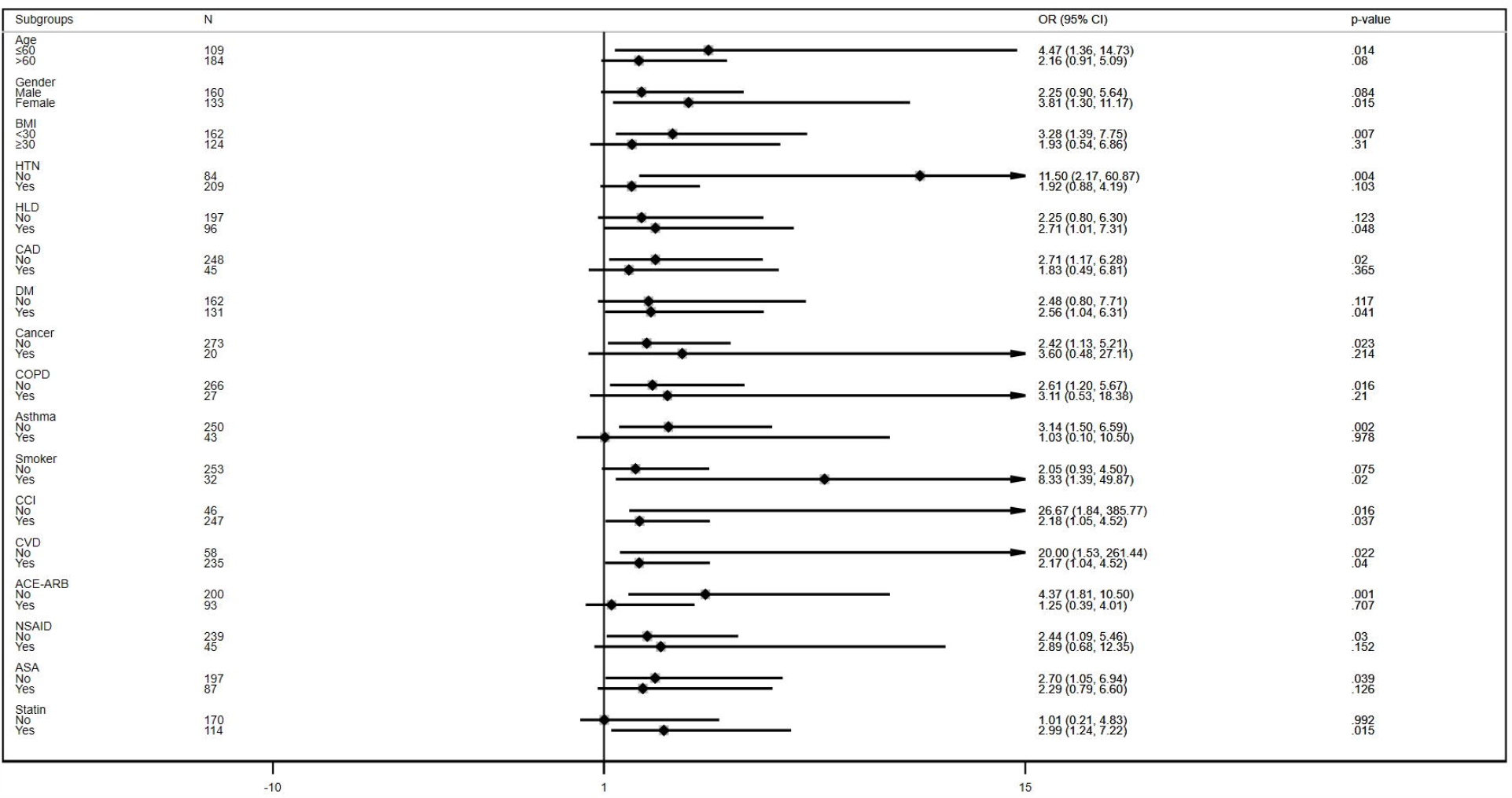
Forest Plot showing the effects of PPI in each subgroup. Forest Plot indicates that PPI increased the odds of mortality regardless of any patient subgroups. It also shows that PPI increased the odds of mortality in patients who had CCI (at least 1 or higher) or received statin. Even patients without having cardiac risk factors (DM, HTN, HLD, and CAD) or CCI of zero had a higher odds of mortality if they received PPI. PPI did not increase the odds of mortality only among patients who had asthma, received ACE-ARB, and did not receive a statin. Footnotes: BMI: Body mass index; HTN: Hypertension; HLD: Hyperlipidemia; CAD: Coronary artery disease; DM: Diabetes mellitus; COPD: Chronic obstructive pulmonary disease; CCI: Charlson’s comorbidity index; CVD: Cardiovascular disease; ACE-ARB: Angiotensin-converting enzymes inhibitors-angiotensin II receptor blockers; NSAIDs: Nonsteroidal anti-inflammatory drugs; ASA: Aspirin; OR: odds ratio.

## Discussion

In our study, we found that 15.6% of hospitalized COVID-19 patients were on PPIs at home. PPI-users had more than two-fold higher risks of mortality and mechanical ventilation requirement than non-users, even after adjusting for potential confounding variables. Our subgroup analyses showed that PPI users had markedly worse prognosis in patients without underlying cardiac comorbidities.

PPIs predominately exert their effect by reducing gastric acid output via irreversible proton pump inhibition [10]. This profound hypochlorhydria can diminish the protective effect of gastric acid, which can theoretically increase the survival of SARS-CoV-2 in the stomach and invading the GI epithelial cells [1, 8, 11]. In the study by Xiao et al., viral host receptor ACE2 and viral nucleocapsid protein stained positive in the cytoplasm of GI epithelial cells suggesting that the SARS-CoV-2 virus invades the mucosal cells, multiply and produce virions [1]. In addition, the reduced acid output due to PPI can change the composition of the gut microbiome, which could increase enteric infections [9, 12, 13]. Furthermore, digestive symptoms such as altered taste, nausea, loss of appetite, and diarrhea are common in COVID-19 patients, and few studies reported worse outcomes with the presence of GI symptoms [2, 5]. In our study, the PPI group did not have higher GI symptoms compared to controls. It remains to be studied in prospective studies if individuals with long-term PPI users are more susceptible to manifest GI symptoms.

Although in our study, the PPI-group is associated with increased mortality after adjustment of potential confounders, these differences should be interpreted with caution as there could be potential unrecognizable confounders. The biological plausibility of such association needs further elaboration, especially with changes in acid secretion and viral transmission through the GI tract. Further studies are urgently needed to evaluate the effect of pH changes in the infectivity of SARS-CoV-2. Regardless, these findings are vital and provide a roadmap for future studies to evaluate the effect of PPI use and outcomes in COVID-19.

Important limitations exist for this study. Our study is a single-center, retrospective, small-sized study which could introduce bias and limit its generalizability. Given the retrospective nature of the study, we could only include the information which was available in the medical records. We were unable to determine the information about the type, dose, duration, frequency, and compliance for the PPI use since we did not have access to the outpatient medical records. Lastly, potential unrecognized confounders could have been missed in our study. Despite these limitations, the main strength of the study is that it addresses a clinically significant issue about the implication of pre-hospitalization PPI use and COVID-19.

In conclusion, Pre-hospitalization PPI use is prevalent in COVID-19 infected patients and associated with increased mortality and MV support among hospitalized COVID-19 patients. The results of our study warrant further investigation to evaluate if PPI-induced acid suppression is associated with worse clinical outcomes primarily because of the omnipresence of ACE-2 in the GI tract.

## Data Availability

Not available

## Acknowledgments

None

